# The α-Synuclein seeding assay discriminates between LRRK2 p.Gly2019Ser variant carriers with and without Parkinson’s disease

**DOI:** 10.64898/2026.05.13.26353087

**Authors:** Theresa Lüth, Carolin Gabbert, Teresa Kleinz, Christoph Much, Björn-Hergen Laabs, Sebastian Sendel, Inke R. König, Amke Caliebe, Matthew Farrer, Brian Fiske, Cornelis Blauwendraat, Christine Klein, Joanne Trinh, Global Parkinson’s Genetics Program (GP2)

## Abstract

**Background:** Reliable biomarkers for Parkinson’s disease (PD) pathology detection are essential for research. The α-synuclein (aSyn) seed amplification assay (SAA) is a validated biomarker for misfolded aSyn.

**Objectives:** To assess the association between aSyn SAA and *LRRK2*-related PD (*LRRK2*-PD) and its link to mitochondrial genetic burden.

**Methods:** We included N=76 LRRK2 p.Gly2019Ser variant carriers (N=22 affected, N=54 unaffected), N=714 patients with idiopathic PD (iPD), and N=411 controls from Norway. We analyzed cerebrospinal fluid (CSF)-based aSyn SAA in N=10 PD patients and N=30 unaffected *LRRK2* p.Gly2019Ser carriers, alongside N=6 controls and N=56 iPD patients. A mitochondrial polygenic score (MGS) was derived from genotyping data, using PPMI as an additional cohort (iPD: N=355, *LRRK2*-PD: N=118).

**Results:** Seeding was observed in 80%of patients with *LRRK2*-PD, and in one unaffected variant carrier (AUC=0.97, CI 0.92-1.00). In a meta-analysis across two PD cohorts, higher MGS was associated with increased aSyn seeding (pooled β=0.38, p=0.028).

**Conclusions:** CSF-based aSyn SAA can discriminate between *LRRK2*-PD and unaffected carriers. Our findings support an association with mitochondrial burden and aSyn seeding.

## Introduction

The early diagnosis of individuals affected with Parkinson’s disease (PD) is critical to ensure effective treatment, symptom management, and identification of potential participants for clinical trials. Thus, a biomarker for accurately predicting disease onset and progression and for identifying high-risk individuals is of immense importance.

A recently established biomarker assay for PD is the α-Synuclein (aSyn) seed amplification assay (SAA). It has been demonstrated to discriminate between PD patients and healthy individuals accurately^1, 2^. One of the most extensive studies evaluating aSyn SAA reported positive seeding in 4%of healthy individuals and in ∼98%of patients with sporadic PD;however, the aSyn SAA positivity was significantly lower for patients with *LRRK2*-related PD (*LRRK2*-PD) at approximately 67%^3^. The substantially lower seeding rate for particularly *LRRK2*-related or *PRKN/PINK1*-related PD remains elusive but might reflect underlying biological differences in the pathogenesis of different PD subtypes. Indeed, aSyn aggregation, particularly in the *substantia nigra*, is widely considered a pathological hallmark of PD, but not all patients exhibit the classical aSyn aggregates at *post-mortem*. In addition, aSyn aggregates have been observed in other disorders, including neurodegeneration with brain iron accumulation, sclerosing panencephalitis, dementia with Lewy bodies, and Alzheimer’s disease. Thus, while Lewy bodies are an important feature of PD and are included in the diagnostic criteria, they are not entirely specific and may not be present in all cases. Additional biomarkers (e.g., mitochondrial polygenic burden or heteroplasmic variant load) might enhance the earlier detection of PD by capturing a broader range of biological pathways implicated in PD pathogenesis and may improve PD subtyping.

Recently, the importance of mitochondrial dysfunction has also been observed in alpha-synuclein-related PD (SNCA-PD)^4^. Beyond the rare monogenic SNCA-related PD form, accumulation of aSyn aggregates also impairs mitochondrial complex I activity and increases ROS, leading to membrane permeabilization, neurotoxicity, and cell death. This highlights the convergence of mitochondrial dysfunction and α-syn aggregation in PD pathogenesis.

Mitochondrial genetic burden, as summarized by a mitochondrial polygenic score (MGS), has also been shown to be associated with PD affection status, including *LRRK2*-PD^5, 6^. Herein, we aimed to assess the association between aSyn SAA and *LRRK2*-PD, and its link to mitochondrial genetic burden.

## Methods

### Norwegian study cohort and alpha-synuclein seed amplification assay

We included N=76 *LRRK2* p.Gly2019Ser variant carriers, of whom N=22 were affected, and 54 were not affected with PD. The *LRRK2* p.Gly2019Ser variant was confirmed by Sanger sequencing. The mean age at examination (AAE) was 70.7 years among the affected and 60.8 years among the unaffected, and the mean age at onset (AAO) was 57.0 years (**Table 1**). Additionally, we included 714 patients with idiopathic PD (iPD) and 411 healthy controls with a mean AAE of 73.4 years and 62.6 years, respectively. All participants were recruited from Norway, referred by general practitioners or other hospitals, and underwent clinical examination and longitudinal follow-up at outpatient clinics across three hospitals in Central Norway.

**Table 1.** Demographic overview and outcome of the α-Synuclein seed amplification assay in a Norwegian cohort of LRRK2 p.Gly2019Ser variant carriers.

| Demographics |  |  |  |  |  |  |
| --- | --- | --- | --- | --- | --- | --- |
|  | N<br>(N<sub>CSF</sub>) | N of men<br>(%) | Mean AAE<br>(±SD) | Mean AAO<br>(±SD) | N of<br>positive<br>aSyn SAA | N of<br>negative<br>aSyn SAA |
| Affected *LRRK2*<br>variant carriers | 22<br>(10) | 11 (50%) | 70.7 years<br>(±14.5) | 57.0<br>(±12.9) | 8 | 2 |
| Unaffected *LRRK2*<br>variant carriers | 54<br>(30) | 38 (46.9%) | 60.8 years<br>(±15.3) | NA | 1 | 29 |
| Idiopathic PD | 714<br>(56) | 469 (65.7%) | 73.4 years<br>(±9.4) | 60.4 years<br>(±10.1) | 54 | 2 |
| Healthy controls | 411<br>(6) | 223 (54.3%) | 62.6 years<br>(±11.5) | NA | 0 | 6 |
| Association between aSyn SAA and mitochondrial polygenic score in patients with PD from the Norwegian population* |  |  |  |  |  |  |
|  | Estimate |  | Standard error |  | p-value |  |
| MGS<sub>std</sub> | 0.71 |  | 0.60 |  | 0.236 |  |
| PD-subtype: iPD | 1.52 |  | 1.23 |  | 0.217 |  |
| AAO | 0.02 |  | 0.05 |  | 0.587 |  |
| Sex: Women | -0.86 |  | 1.32 |  | 0.517 |  |
| Association between aSyn SAA and mitochondrial polygenic score in patients with PD from PPMI* |  |  |  |  |  |  |
|  | Estimate |  | Standard error |  | p-value |  |
| MGS<sub>std</sub> | 0.35 |  | 0.18 |  | 0.046 |  |
| PD-subtype: iPD | 2.84 |  | 0.36 |  | 4.2×10<sup>-15</sup> |  |
| AAO | -0.10 |  | 0.02 |  | 5.1×10<sup>-6</sup> |  |
| Sex: Women | -0.57 |  | 0.33 |  | 0.082 |  |

N=Number of participants, N<sub>CSF</sub>=Number of participants with available CSF analyzed with aSyn SAA, AAE=Age at examination, AAO=Age at onset, aSyn SAA=Alpha-synuclein seed amplification assay, NA=Not applicable, PPMI=Parkinson's Progression Markers Initiative, SD=Standard deviation.
\*R formular: glm(formula = aSyn SAA (positive/negative) ~ MGS<sub>std</sub> + PD-subtype (*LRRK2*-PD/iPD) + AAO + Sex, family = binomial)

CSF samples of N=10 patients with *LRRK2*-PD and N=30 unaffected *LRRK2* variant carriers, alongside N=6 healthy controls and N=56 patients with iPD were analyzed using aSyn SAA. All patients with *LRRK2*-PD and 44 of the iPD patients had DNA available for genetic testing.

The aSyn SAA was performed at Amprion Inc. according to an established protocol^7^. In the assay, the aSyn seeds undergo cyclic amplification, involving fragmentation into self-propagating units, which is followed by elongation using recombinant aSyn as a substrate. Subsequently, the fluorescence is detected using a dye specific to the resulting fibrils (i.e., thioflavin T)^7^.

### Genetic data and mitochondrial polygenic score

Two genotyping arrays were used in this study: the NeuroBooster array (NBA, v1.0, Illumina, San Diego, CA) and the Infinium Global Screening Array with custom content for neurodegenerative-related variants (GSA). NBA genotyping was performed at GP2 (Release 10), and GSA was performed at the Institute of Clinical Molecular Biology, Kiel. NBA genotyping data were processed by GenoTools^8^ and imputed using the TOPMed reference panel. Further information on GP2 data acquisition, imputation, quality control, and release policies is available at https://gp2.org/.

The *LRRK2* variant carriers were genotyped separately utilizing the GSA, assessing ∼700,000 variants. The genotyping data obtained from the GSA array were imputed using the Michigan Imputation Server (v2, Reference Panel: HRC r1.1, Rsq filter: 0.3) as previously described^5^.

After the imputation, both genotyping datasets were quality controlled analogously (minor allele frequency >0.01, missingness per sample <0.02, missingness per SNP <0.05, and Hardy-Weinberg equilibrium p-value >1x10^-50^).

Lastly, whole-genome sequencing data from the (Parkinson’s Progression Markers Initiative) (PPMI) cohort were quality controlled analogously. We included N=355 iPD patients (mean AAO (±SD)=61.3 years (±9.65), men: N=235 (66%), positive aSyn SAA: N=339 (95%)) and N=118 LRRK2-PD patients ((mean AAO (±SD)=60.5 years (±8.97), men: N=66 (55%), positive aSyn SAA: N=76 (64%)).

The MGS was calculated with *PLINK* (version 1.9)^9^ score function utilizing our published ∼15,000 SNPs located in mitochondrial function-associated genes and corresponding weights^5^ (https://github.com/LuethTheresa/MitochondrialPolygenicScoreAndAgeAtOnset). The obtained MGS was standardized to a mean of 0 and a standard deviation (SD) of 1 across the entire cohort.

### Statistical analysis

Statistical analyses were performed with R (version 4.3.1)^10^. All analyses in this study were exploratory, and p-values cannot be interpreted for significance. The sensitivity and specificity of the aSyn SAA, together with age at examination and sex, were evaluated using ROC analysis based on a logistic regression model implemented with the *pROC* R package^11^. As a proof of concept, the association between iPD affection status and MGS was assessed in the Norwegian cohort using logistic regression, adjusting for sex, age, and principal components 1-2. Associations between MGS and aSyn SAA in PD patients were analyzed using multiple logistic regression models adjusted for *LRRK2*-PD/iPD status, sex, and AAO. Fixed-effect meta-analysis was performed across the Norwegian cohort and PPMI using the metafor R package^12^.

### Data Sharing

Data used in the preparation of this article were obtained from the Global Parkinson’s Genetics Program (GP2;https://gp2.org). Specifically, we used Tier 2 data from GP2 release 10 (https://zenodo.org/records/15748014). Tier 1 data can be accessed by completing a form on the Accelerating Medicines Partnership in Parkinson’s Disease (AMP®-PD) website (https://amp-pd.org/register-for-amp-pd). Tier 2 data access requires approval and a Data Use Agreement signed by your institution. Qualified researchers are encouraged to apply for direct access to the data through AMP-PD.

Data used in the preparation of this article were obtained on 01/04/2025 from the Parkinson’s Progression Markers Initiative (PPMI) database (RRID:SCR_006431). For up-to-date information on the study, visit www.ppmi-info.org.

Genotyping data for the *LRRK2* variant carriers generated within DFG FOR2488 included in this study, as well as α-synuclein seed amplification assay outcomes, are not currently available through GP2 but will be incorporated in future releases. In the meantime, these data are available to qualified researchers upon reasonable request to the corresponding author. All code generated for this article, and the identifiers for all software programs and packages used, are available on GitHub [https://github.com/LuethTheresa/MitochondrialPolygenicScoreAndAgeAtOnset] and were given a persistent identifier via Zenodo [DOI: 10.5281/zenodo.19664620].

## Results

### aSyn SAA in *LRRK2*-PD and iPD

The aSyn SAA was positive in eight out of ten patients with *LRRK2*-PD (80%). The aSyn SAA was also positive for one out of 30 (3%) unaffected individuals with the *LRRK2* p.Gly2019Ser variant (age at sample collection range=60-70 years). Utilizing a logistic regression model with affection status as outcome and the aSyn SAA as a biomarker, plus AAE and sex as influence variables, the ROC curves and respective AUC values indicate very good discrimination between affected and unaffected *LRRK2* p.Gly2019Ser variant carriers (AUC=0.97, CI 0.92–1.00, **Supplementary Figure 1**). Assessing the individual prediction capability of the aSyn SAA outcome, without sex and age as covariates, the AUC still indicated good discrimination (AUC=0.883, CI=0.75-1.00).

However, the fraction of positive aSyn SAA was lower in the *LRRK2*-PD group compared to the iPD group. In the Norwegian cohort, 54 out of 56 patients with iPD showed a positive aSyn SAA (96%), and none of the healthy controls had a positive aSyn SAA (**Table 1**).

### Assessing the relationship between aSyn seeding and mitochondrial genetic burden

We included the MGS reflecting the mitochondrial function-associated genetic burden. We have previously shown that a higher MGS is associated with PD affection status^5, 6^. As proof of concept, we assessed the association of MGS with iPD (N=714) and healthy controls (N=411) from the Norwegian cohort. The MGS was higher among patients with iPD compared to healthy controls (**Supplementary Figure 2**), and applying a linear multiple regression model showed an association between MGS and iPD affection status (β=0.17, SE=0.07, p=0.02). Next, we explored the association between aSyn SAA outcome and MGS in patients with PD who had available aSyn SAA seeding and genotyping data. In a logistic regression (iPD with aSyn positive seeding: N=42/44;*LRRK2*-PD with aSyn positive seeding: N=8/10), higher MGS was associated with increased odds of positive aSyn SAA;however, the effect estimate was imprecise, likely reflecting limited sample size (β=0.71, SE=0.24, p=0.236;**Table 1**). Given our limited sample size and low statistical power, we also utilized the genetic and aSyn SAA data from the PPMI cohort. Applying the same multiple logistic regression model, we observed a similar association between higher MGS and positive SAA (iPD: N=355, *LRRK2*-PD: N=118;β=0.35, p=0.046, **Table 1**). Across the Norwegian and PPMI cohorts, a fixed-effects meta-analysis showed that higher MGS was significantly associated with increased aSyn seeding (pooled β=0.38, OR=1.46, p=0.028, **Supplementary Figure 3**), indicating that a one SD increase in MGS is associated with a 46%higher odds of aSyn seeding. To assess whether the association between increased MGS and aSyn seeding was specific to mitochondrial polygenic burden, we calculated the Nalls *et al*. PD polygenic risk score (PRS)^13^, which is not related to a particular pathway. None of the ∼15,000 variants included in the MGS overlap with the 90 variants comprising the Nalls PRS. There was no association between the PGS and aSyn seeding outcome (β=0.225, p=0.271) in PPMI.

## Discussion

Our results might not yet indicate the clinical applicability of CSF-based aSyn SAA to determine the affection status of *LRRK2* p.Gly2019Ser variant carriers, but highlight its importance as a research tool to explore biological insights of PD pathogenesis. A literature review on aSyn SAA and *LRRK2*-related PD showed a detection rate ranging from 38%to 79%^3, 14-19^ among patients with *LRRK2*-PD (**Supplementary Text** 1, Supplementary Table 1). As we observed aSyn seeding in 80%of patients with *LRRK2*-PD, the SAA results were well in line with previously published studies. The assessment of CSF-based aSyn SAA results from unaffected *LRRK2* variant carriers is understudied;however, the screening of the PPMI cohort reported that 14/159 (9%) unaffected *LRRK2* variant carriers showed aSyn seeding^3^. A recent study on *LRRK2* p.Gly2019Ser variant carriers of Ashkenazi Jewish descent reported a seeding rate of 0/11 (0%) in unaffected variant carriers^20^. In our small Norwegian cohort, one of the CSF samples from unaffected *LRRK2* variant carriers (3%) showed aSyn seeding, whereas none of the CSF samples from healthy controls did (0%). We observed a higher rate of seeding for unaffected *LRRK2* carriers compared to controls, though the number of controls in our cohort is small for a fair comparison. There is an anticipated higher positivity rate in carriers compared to healthy controls, as reflected by previous studies^3^.

The higher variability in aSyn SAA results among patients with *LRRK2*-PD may reflect biological differences in the underlying molecular mechanisms driving PD pathogenesis compared to patients with iPD. Additionally, the group of *LRRK2* variant carriers in study cohorts is usually smaller and might be further impacted by ancestry or ethnicity differences, possibly contributing to the observed variability. Interestingly, mitochondrial forms of monogenic PD, particularly PRKN-PD, are less frequently associated with aSyn deposition or Lewy bodies^3^. However, it has been shown that patients with *LRRK2*-PD without Lewy body pathology still show aSyn neuronal aggregation^21, 22^. Thus, identifying factors associated with aSyn SAA outcome in variant carriers is essential to improve the overall predictive accuracy of PD affection status and PD subtyping, which may require assessing multiple biomarkers.

It has been reported that a higher PD polygenic score^13^ was associated with aSyn seeding in a cohort of eleven patients with *LRRK2*-PD, but this finding was not replicated in the larger PPMI cohort^15^. Given the importance of mitochondrial dysfunction in (*LRRK2*-related) PD and the recently demonstrated direct connection between aSyn aggregation and mitochondrial damage^4^, we assessed the relationship between an MGS and aSyn SAA results. Together, our findings suggest an association between higher mitochondrial genetic burden and positive aSyn seeding, underscoring the value of integrating multiple biomarkers in PD. This relationship suggests a potential biological convergence between mitochondrial dysfunction and aSyn aggregation in PD pathogenesis. Interestingly, aSyn seeding showed no association with the general PD PGS^13^, supporting the notion that its association with the MGS might be specific to mitochondrial dysfunction. Importantly, none of the specific ∼15,000 variants included in the MGS overlap with the 90 variants comprising the Nalls PRS, indicating that these genetic measures capture distinct components of PD risk. Future studies with larger cohorts should further explore mitochondrial impairment and aSyn aggregation across *LRRK2*-PD and other PD subtypes.

## Data Availability

Data used in the preparation of this article were obtained from the Global Parkinson's Genetics Program (GP2;https://gp2.org). Specifically, we used Tier 2 data from GP2 release 10 (https://zenodo.org/records/15748014). Tier 1 data can be accessed by completing a form on the Accelerating Medicines Partnership in Parkinson's Disease (AMP-PD) website (https://amp-pd.org/register-for-amp-pd). Tier 2 data access requires approval and a Data Use Agreement signed by your institution. Qualified researchers are encouraged to apply for direct access to the data through AMP-PD.
Data used in the preparation of this article were obtained on 01/04/2025 from the Parkinson's Progression Markers Initiative (PPMI) database (RRID:SCR_006431). For up-to-date information on the study, visit www.ppmi-info.org.
Genotyping data for the LRRK2 variant carriers generated within DFG FOR2488 included in this study, as well as α-synuclein seed amplification assay outcomes, are not currently available through GP2 but will be incorporated in future releases. In the meantime, these data are available to qualified researchers upon reasonable request to the corresponding author. All code generated for this article, and the identifiers for all software programs and packages used, are available on GitHub [https://github.com/LuethTheresa/MitochondrialPolygenicScoreAndAgeAtOnset] and were given a persistent identifier via Zenodo [DOI: 10.5281/zenodo.19664620].
 

## Acknowledgment

The authors wish to acknowledge the late Professor Jan Aasly, for his expertise in movement disorders, the collaboration, resourcefulness, and generosity. He was responsible for the collection of samples and the assessment of the Norwegian participants analyzed in this study. We are grateful for his contributions, which made this work possible.

PPMI –a public-private partnership –is funded by the Michael J. Fox Foundation for Parkinson’s Research and funding partners, including 4D Pharma, Abbvie, AcureX, Allergan, Amathus Therapeutics, Aligning Science Across Parkinson’s, AskBio, Avid Radiopharmaceuticals, BIAL, BioArctic, Biogen, Biohaven, BioLegend, BlueRock Therapeutics, Bristol-Myers Squibb, Calico Labs, Capsida Biotherapeutics, Celgene, Cerevel Therapeutics, Coave Therapeutics, DaCapo Brainscience, Denali, Edmond J. Safra Foundation, Eli Lilly, Gain Therapeutics, GE HealthCare, Genentech, GSK, Golub Capital, Handl Therapeutics, Insitro, Jazz Pharmaceuticals, Johnson &Johnson Innovative Medicine, Lundbeck, Merck, Meso Scale Discovery, Mission Therapeutics, Neurocrine Biosciences, Neuron23, Neuropore, Pfizer, Piramal, Prevail Therapeutics, Roche, Sanofi, Servier, Sun Pharma Advanced Research Company, Takeda, Teva, UCB, Vanqua Bio, Verily, Voyager Therapeutics, the Weston Family Foundation and Yumanity Therapeutics.

This project was supported by the Global Parkinson’s Genetics Program (GP2) (https://gp2.org). GP2 is funded by the Aligning Science Across Parkinson’s (ASAP) (https://ror.org/03zj4c476) initiative and implemented by The Michael J. Fox Foundation for Parkinson’s Research (https://ror.org/03arq3225). For a complete list of GP2 members, see https://doi.org/10.5281/zenodo.7904831.

## Authors’Roles

(1) Research Project: A. Conception and Design, B. Data Acquisition, C. Data Analysis;(2) Statistical Analysis: A. Design, B. Execution, C. Review and Critique;(3) Manuscript Preparation: A. Drafting the Manuscript and/or Figures, B. Review and Critique.

Theresa Lüth: 1A,B,C;2A,B;3A

Carolin Gabbert: 1B, C;2C;3B

Teresa Kleinz: 1B, C;2C;3B

Christoph Much: 1B;3B

Björn-Hergen Laabs: 2A, C;3B

Sebastian Sendel: 2A, C;3B

Inke R. König: 2A, C;3B

Amke Caliebe: 2A, C;3B

Matthew Farrer: 1B;2C;3B

Brian Fiske: 1A,B;2C;3B

Cornelis Blauwendraat: 1B;2C;3B

Christine Klein: 1A,B;2C;3A,B

Joanne Trinh: 1A,B;2A,C;3A,B

## Financial Disclosures of all authors (for the preceding 12 months)

**Financial Disclosures of all authors (for the preceding 12 months):**
| **Autor** | **Disclosures** |
| --- | --- |
| TL | Nothing to declare |
| CG | Nothing to declare |
| TK | Nothing to declare |
| CM | Nothing to declare |
| BHL | Nothing to declare |
| SS | Nothing to declare |
| IRK | Nothing to declare |
| AC | Nothing to declare |
| MF | Nothing to declare |
| BF | Nothing to declare |
| CB | Aligning Science Across Parkinson's (ASAP) Global Parkinson's Genetics Program (GP2) |
| CK | CK has served as medical advisor to Centogene, and Bial. She received speakers' honoraria from Bial and royalties from Oxford University Press and Springer Nature. Aligning Science Across Parkinson's (ASAP) Global Parkinson's Genetics Program (GP2). |
| JT | JT is supported by the German Research Foundation (DFG), Aligning Science Across Parkinson's (ASAP) Global Parkinson's Genetics Program (GP2), is a consultant for Acurex and on the advisory board for Synlab international. She receives royalties from Elsevier and a speaker's honorarium from the EAN. |

## Ethics statement

The details of the IRB/oversight body that provided approval or exemption for the research described are given below:

1. Written informed consent was obtained from all individuals and approved by the Ethics Committee at the University of Lübeck, Lübeck, Germany.
2. Data used in the preparation of this article were obtained from the Global Parkinson’s Genetics Program (GP2;https://gp2.org). Specifically, we used Tier 2 data from GP2 release 10 (https://zenodo.org/records/15748014). Tier 1 data can be accessed by completing a form on the Accelerating Medicines Partnership in Parkinson’s Disease (AMP®-PD) website (https://amp-pd.org/register-for-amp-pd). Tier 2 data access requires approval and a Data Use Agreement signed by your institution. Qualified researchers are encouraged to apply for direct access to the data through AMP-PD.
3. The PPMI study was conducted in accordance with the Declaration of Helsinki and the Good Clinical Practice (GCP) guidelines after approval of the local ethics committees of the participating sites.

