## Supplementary for "The α-Synuclein seeding assay discriminates between LRRK2 p.Gly2019Ser variant carriers with and without Parkinson’s disease"

**Supplementary Text 1.** **Literature review on *LRRK2-*related PD and α-Synuclein seed amplification assay.**

We performed a literature review on the performance of α-Synuclein seed amplification assay (aSyn SAA) in *LRRK2*-related Parkinson's disease (PD). We utilized the following search term to screen PubMed:

("Alpha-synuclein seeding amplification" OR "Alpha-synuclein seeding amplification assay" OR "α-synuclein seeding amplification" OR "α-synuclein amplification" OR "α-synuclein seeding amplification assay" OR "a-synuclein seeding amplification" OR "a-synuclein amplification" OR "a-synuclein seeding amplification assay" OR "Alpha-Synuclein Seed Amplification Assay" OR "α-Synuclein Seed Amplification Assay" OR "a-Synuclein Seed Amplification Assay" OR "αSyn-SAA" OR "αSyn SAA" OR "aSyn-SAA" OR "aSyn SAA") AND (LRRK2 OR PARK8 OR "Leucine-rich repeat kinase 2") AND ("english"[Language])

The screening resulted in 18 publications. We excluded papers published before 2020 (N=5), reviews (N=1), pre-prints that have also been published in peer-reviewed journals afterwards (N=1) and publications that did not report aSyn SAA on LRRK2 variant carriers (N=2). Thus, we include N=9 publications, and N=7 studies reported the results of aSyn SAA in *LRRK2*-related PD. The results are summarized in **Supplementary Table 1**.

**Supplemental Table 1. Overview of studies about the performance of the α-Synuclein seed amplification assay (aSyn SAA) in *LRRK2*-related Parkinson's disease (PD).**

| **Ref** | **Summary** | ***LRRK2*-related aSyn SAA results reported** |
| --- | --- | --- |
| Schumacher JG, Zhang X, Macklin EA, et al. Baseline α-synuclein seeding activity and disease progression in sporadic and genetic Parkinson's disease in the PPMI cohort. Preprint. medRxiv. 2025;2024.09.27.24311107. Published 2025 Apr 2. doi:10.1101/2024.09.27.24311107 | Baseline aSyn SAA data were assessed from 564 PPMI participants (including N=162 *LRRK2*-PD), and no statistically significant associations were observed between baseline α-syn seeding activity and PD progression among patients. | Not reported |
| Chahine LM, Lafontant DE, Choi SH, et al. LRRK2-associated parkinsonism with and without in vivo evidence of alpha-synuclein aggregates: longitudinal clinical and biomarker characterization. Brain Commun. 2025;7(2):fcaf103. Published 2025 Mar 6. doi:10.1093/braincomms/fcaf103 | aSyn SAA data from PPMI participants (including N=148 *LRRK2*-PD) were assessed, and *LRRK2*-PD patients without seeding exhibit less severe motor manifestations and decline. | A subset of the PPMI cohort has been evaluated, reporting aSyn seeding in  68% (102/148) of the samples |
| Goldstein O, Shani S, Gana-Weisz M, et al. The effect of polygenic risk score on PD risk and phenotype in LRRK2 G2019S and GBA1 carriers. J Parkinsons Dis. 2025;15(2):291-299. doi:10.1177/1877718X241310722 | Genotyping of 786 patients with PD was conducted, along with aSyn SAA, on a subset of samples. *LRRK2*-PD patients with positive aSyn SAA had a higher PRS compared to those with a negative aSyn SAA. | 46% (6/13) |
| Droby A, Yoffe-Vasiliev A, Atias D, et al. Radiological markers of CSF α-synuclein aggregation in Parkinson's disease patients. NPJ Parkinsons Dis. 2025;11(1):7. Published 2025 Jan 3. doi:10.1038/s41531-024-00854-4 | Assessment of CSF radiological measures in 41 PD patients (including N=13 *LRRK2*-PD) and 14 healthy controls showed that PD-SAA+ status was associated with reduced regional brain volumes, altered caudal FC, and lower SBRs, whereas these changes were less pronounced in PD-SAA-, potentially indicating a milder neurodegenerative process. | 38% (5/13) |
| Grillo P, Concha-Marambio L, Pisani A, Riboldi GM, Kang UJ. Association between the Amplification Parameters of the α-Synuclein Seed Amplification Assay and Clinical and Genetic Subtypes of Parkinson's Disease. Mov Disord. 2025;40(2):305-314. doi:10.1002/mds.30085 | Assessment of clinical and CSF-αSyn-SAA data from PPMI. CSF-αSyn-SAA was positive in most PD cases (*LRRK2*-PD: 77%, *GBA*1-PD: 92.3%, sporadic PD: 93.8%), with amplification parameters varying across subtypes; notably, *LRRK2*-PD showed longer T50/TTT and smaller AUC compared to *GBA*-PD and sporadic PD. | A subset of the PPMI cohort has been evaluated, reporting aSyn seeding in  77% (95/124) of the samples |
| Venuto CS, Herbst K, Chahine LM, Kieburtz K. Predicting Cerebrospinal Fluid Alpha-Synuclein Seed Amplification Assay Status from Demographics and Clinical Data. Preprint. medRxiv. 2025;2024.08.07.24311578. Published 2025 Mar 7. doi:10.1101/2024.08.07.24311578 | PPMI and Systemic Synuclein Sampling Study (S4) data were assessed, showing that data-driven models based on non-invasive clinical features can accurately predict CSF aSyn SAA status. | Not reported |
| Yuan Y, Wang Y, Liu M, et al. Peripheral cutaneous synucleinopathy characteristics in genetic Parkinson's disease. Front Neurol. 2024;15:1404492. Published 2024 May 1. doi:10.3389/fneur.2024.1404492 | This study aimed to detect p-α-syn deposition characteristics in rare genetic PD patients (*CHCHD2* and *RAB39B*) and other genetic forms of PD. A Syn SAA was performed on skin biopsy samples. | 78.57% (11/14) |
| Siderowf A, Concha-Marambio L, Lafontant DE, et al. Assessment of heterogeneity among participants in the Parkinson's Progression Markers Initiative cohort using α-synuclein seed amplification: a cross-sectional study. Lancet Neurol. 2023;22(5):407-417. doi:10.1016/S1474-4422(23)00109-6 | Cross-sectional analysis of CSF-based aSyn SAA in the PPMI cohort. | 67.5% (83/123) |
| Coughlin DG, Shifflett B, Farris CM, et al. α-Synuclein Seed Amplification Assay Amplification Parameters and the Risk of Progression in Prodromal Parkinson Disease. Neurology. 2025;104(5):e210279. doi:10.1212/WNL.0000000000210279 | CSF-based aSyn SAA from PPMI was assessed, and positivity and amplification parameters across NMCs, prodromal PD, and PD were compared. αSyn-SAA+ participants with prodromal PD and PD showed faster TTT and T50 and higher AUC than αSyn-SAA+ NMCs and HCs. | A subset of the PPMI cohort has been evaluated, reporting aSyn seeding in 68% (76/112) of the samples |

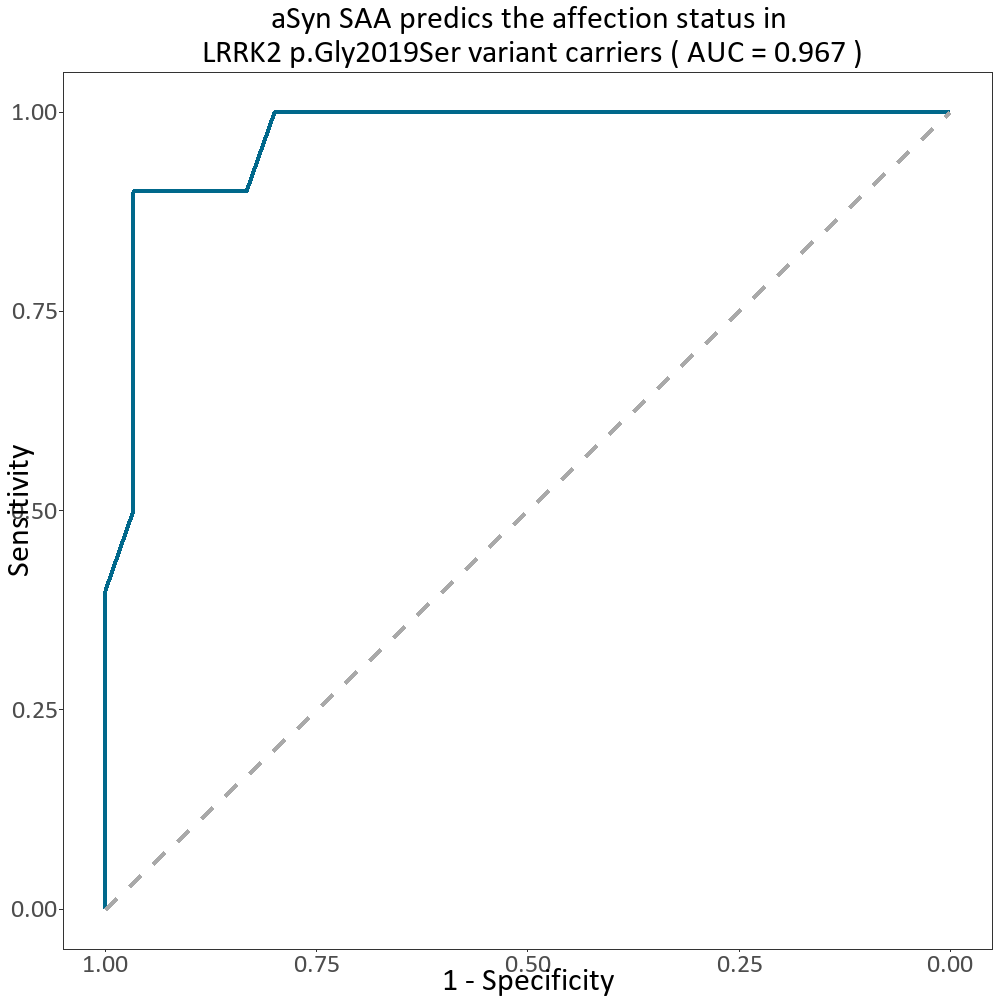

**Supplementary Figure 1. Accuracy of α-Synuclein seed amplification assay (aSyn SAA) to predict the affection status in a cohort of LRRK2 p.Gly2019Ser variant carriers.** The receiver operating characteristic (ROC) curves and respective area under the curve values are displayed. The AUC indicated good discrimination for affected (N=10) versus unaffected (N=29) of LRRK2 p.Gly2019Ser variant carriers, utilizing a logistic regression model with affection status as outcome and the aSyn SAA as a biomarker, plus AAE and sex as influence variables (AUC = 0.97, CI 0.92–1.00).

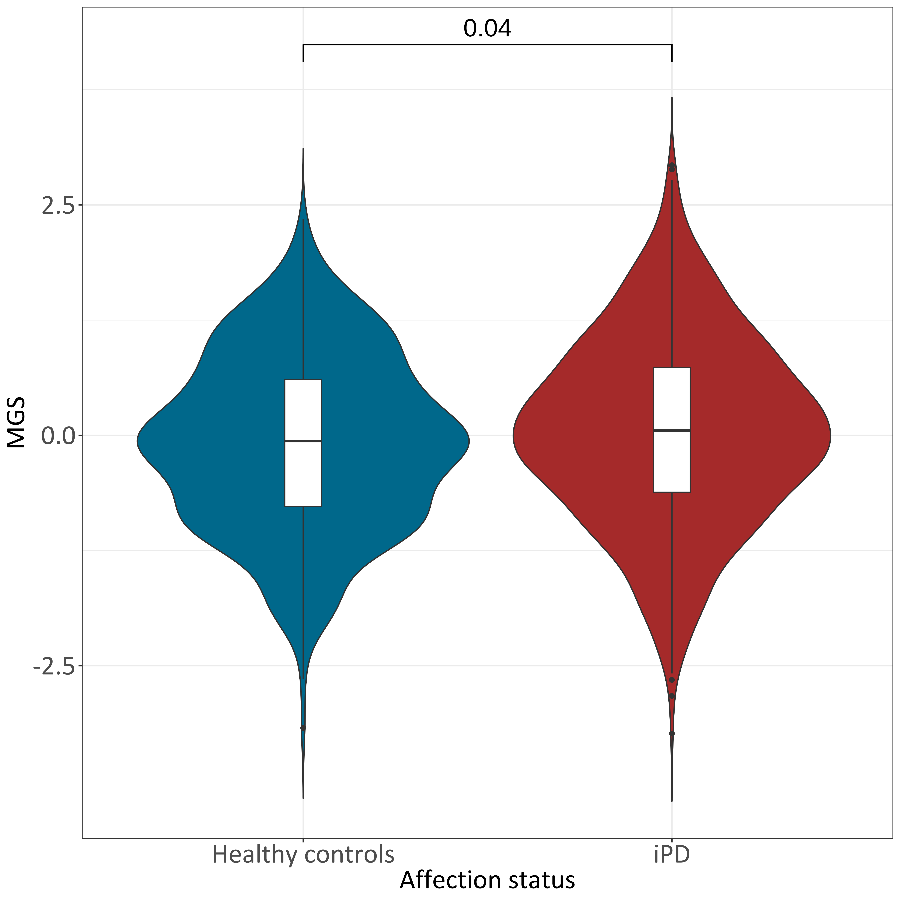

**Supplementary Figure 2.** Associations between mitochondrial polygenic scores (MGS) and idiopathic Parkinson's disease (iPD) status. Violin plots showing the distribution of standardized MGS iPD patients and healthy individuals in the Norwegian population. Boxplots within the violins indicate the median (solid line) and interquartile range (box edges), while the violin shape reflects the overall data distribution. p: t-test p-value.

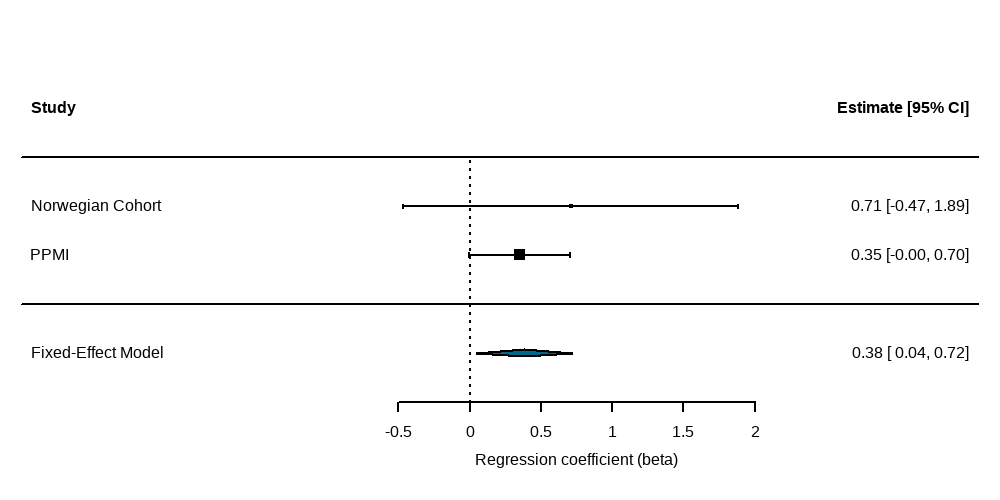

**Supplementary Figure 3. Fixed-effects meta-analysis of the association between mitochondrial genetic score (MGS) and aSyn seeding in Parkinson’s disease.** Forest plot showing cohort-specific and pooled effect estimates from logistic regression models assessing the association between MGS and aSyn seeding assay (SAA) positivity. Squares represent cohort-specific β estimates with horizontal lines indicating 95% confidence intervals (95% CI); square size reflects study weight. The diamond denotes the pooled fixed-effects estimate.
